# Hydroxychloroquine and azithromycin plus zinc vs hydroxychloroquine and azithromycin alone: outcomes in hospitalized COVID-19 patients

**DOI:** 10.1101/2020.05.02.20080036

**Authors:** Philip M. Carlucci, Tania Ahuja, Christopher Petrilli, Harish Rajagopalan, Simon Jones, Joseph Rahimian

## Abstract

**Background:** COVID-19 has rapidly emerged as a pandemic infection that has caused significant mortality and economic losses. Potential therapies and means of prophylaxis against COVID-19 are urgently needed to combat this novel infection. As a result of *in vitro* evidence suggesting zinc sulfate may be efficacious against COVID-19, our hospitals began using zinc sulfate as add-on therapy to hydroxychloroquine and azithromycin. We performed a retrospective observational study to compare hospital outcomes among patients who received hydroxychloroquine and azithromycin plus zinc versus hydroxychloroquine and azithromycin alone.

**Methods:** Data was collected from electronic medical records for all patients being treated with admission dates ranging from March 2, 2020 through April 5, 2020. Initial clinical characteristics on presentation, medications given during the hospitalization, and hospital outcomes were recorded. Patients in the study were excluded if they were treated with other investigational medications.

**Results:** The addition of zinc sulfate did not impact the length of hospitalization, duration of ventilation, or ICU duration. In univariate analyses, zinc sulfate increased the frequency of patients being discharged home, and decreased the need for ventilation, admission to the ICU, and mortality or transfer to hospice for patients who were never admitted to the ICU. After adjusting for the time at which zinc sulfate was added to our protocol, an increased frequency of being discharged home (OR 1.53, 95% CI 1.12-2.09) reduction in mortality or transfer to hospice remained significant (OR 0.449, 95% CI 0.271-0.744).

**Conclusion:** This study provides the first *in vivo* evidence that zinc sulfate in combination with hydroxychloroquine may play a role in therapeutic management for COVID-19.

40-word summary: Zinc sulfate added to hydroxychloroquine and azithromycin may improve outcomes among hospitalized patients.

## INTRODUCTION

The World Health Organization has declared a pandemic due to spread of the coronavirus disease of 2019 (COVID-19) caused by the severe acute respiratory syndrome coronavirus 2 (SARS-CoV2)[1, 2]. SARS-CoV2 is a single-strand RNA coronavirus, which enters human cells mainly by binding the angiotensin converting enzyme 2 (ACE2)[3]. SARS-CoV2 is primarily transmitted after viral particles are inhaled and enter the respiratory tract and has the potential to cause a severe systemic inflammatory response, acute respiratory disease syndrome (ARDS), multi organ failure, and shock[2, 4]. Laboratory abnormalities found in patients with COVID-19 include lymphopenia, elevation in lactate dehydrogenase, C reactive protein, D-dimer, ferritin and interleukin-6 (IL-6)[5, 6].

Several medications are under investigation for the treatment of COVID-19. Despite limited and conflicting data, the U.S. Food and Drug Administration authorized the emergency use of hydroxychloroquine for the treatment of COVID-19 with or without azithromycin. Chloroquine analogues are weak bases that concentrate within acidic endosomes and lysosomes. Once intracellular, chloroquine analogues become protonated and increase pH resulting in prevention of endosomal trafficking, dysfunctional cellular enzymes, and impaired protein synthesis[7]. This inhibits viral replication through interference with endosome-mediated viral entry or late transport of the enveloped virus. Further, this results in interference with the terminal glycosylation of ACE2 receptor expression which prevents SARS-CoV-2 receptor binding and spread of infection [8]. Hydroxychloroquine, a hydroxy-derivative of chloroquine, has also been proposed based on *in vitro* activity against SARS-CoV-2 with a three-fold higher cytotoxic potential compared to chloroquine [9]. However, clinical data in humans has yielded mixed results[10–12]. The anti-viral and anti-inflammatory effects of chloroquine have been suggested to account for its potential utility in preventing COVID-19-related pneumonia. Soon current studies will answer whether hydroxychloroquine is effective as monotherapy or in combination with azithromycin. In the case that hydroxychloroquine is found to be ineffective, it may still have a role to play when combined with zinc sulfate. Zinc inhibits RNA dependent RNA polymerase, and has been shown to do this *in vitro* against SARS-CoV[13]. However, it is difficult to generate substantial intracellular concentrations of zinc, therefore prophylactic administration of zinc alone may not play a role against SarCoV-2[14]. When combined with a zinc ionophore, such as chloroquine (hydroxychloroquine), cellular uptake is increased making it more likely to achieve suitably elevated intracellular concentrations[15]. This combination is already being tested as a prophylactic regimen in a randomized clinical trial.

As New York became the epicenter of the pandemic, hospitals in the area quickly adopted investigational therapies, including the use of hydroxychloroquine and azithromycin. Given this proposed synergistic effect of zinc with hydroxychloroquine, practices at NYULH changed and the addition of zinc sulfate 220 mg PO BID along with hydroxcychloroquine 400 mg once followed by 200 mg PO BID with azithromycin 500 mg once daily became part of the treatment approach for patients admitted to the hospital with COVID-19. This study sought to investigate outcomes among patients who received hydroxychloroquine and azithromycin alone compared to those who received triple therapy with zinc sulfate.

## METHODS

We performed a retrospective analysis of data from patients hospitalized with confirmed SARS-CoV-2 infection at NYU Langone Health. Data was collected from electronic medical records (Epic Systems, Verona, WI) for all patients being treated with admission dates ranging from March 2, 2020 through April 5, 2020. Patients were admitted to any of four acute care NYU Langone Health hospitals across New York City. COVID-19 positivity was determined by real-time reverse-transcriptase-polymerasechain-reaction (RT-PCR) of nasopharyngeal or oropharyngeal swabs. Prior to March 16, tests were completed by the New York City Department of Health and Mental Hygiene. After that date, NYU Langone clinical laboratory conducted tests using the Roche SARS-CoV2 assay in the Cobas 6800 instruments. On March 31, testing was also conducted using the SARS-CoV2 Xpert Xpress assay in the Cepheid GeneXpert instruments. After March 16, only pharyngeal samples were tested.

Patients were included in the study if they were admitted to the hospital, had at least one positive test for COVID-19, received hydroxychloroquine and azithromycin, and had either been discharged from the hospital, transitioned to hospice, or expired. Patients were excluded from the study if they were never admitted to the hospital or if there was an order for other investigational therapies for COVID-19, including tocilizumab, nitazoxanide, rituximab, anakinra, remdesivir, or lopinavir/ritonavir during the course of their hospitalization to avoid potential confounding effects of these medications. We collected demographics as reported by the patient and any past medical history of hypertension, hyperlipidemia, coronary artery disease, heart failure, chronic obstructive pulmonary disease, asthma, malignancy other than non-melanoma skin malignancy, and diabetes. We also recorded vital signs on admission, the first set of laboratory results as continuous variables, and relevant medications as categorical variables, including NSAIDs, anticoagulants, antihypertensive medications and corticosteroids ordered at any point during the course of the hospitalization.

### Statistics

Patients were categorized based on their exposure to hydroxychloroquine (400 mg load followed by 200 mg twice daily for five days) and azithromycin (500 mg once daily) alone or with zinc sulfate (220 mg capsule containing 50 mg elemental zinc twice daily for five days) as treatment in addition to standard supportive care. Descriptive statistics are presented as mean and standard deviation or mean and interquartile range for continuous variables and frequencies for categorical variables. Normality of distribution for continuous variables was assessed by measures of skewness and kurtosis, deeming the dataset appropriate for parametric or nonparametric analysis. A 2-tailed Student’s t test was used for parametric analysis, and a Mann Whitney U test was used for nonparametric data analysis. Pearson’s chi-squared test was used to compare categorical characteristics between the two groups of patients. Linear regression for continuous variables or logistic regression for categorical variables was performed with the presence of zinc as the predictor variable and outcome measures (duration of hospital stay, duration of mechanical ventilation, maximum oxygen flow rate, average oxygen flow rate, average FiO2, maximum FiO2, admission to the intensive care unit (ICU), duration of ICU stay, death/hospice, need for intubation, and discharge destination), as dependent variables. Data was log transformed where appropriate to render the distribution normal for linear regression analysis. Multivariate logistic regression was used to adjust for the timing that our protocol changed to include zinc therapy using admission before or after March 25^th^ as a categorical variable. P-values less than 0.05 were considered to be significant. All analyses were performed using STATA/SE 16.0 software (STATA Corp.).

### Study approval

The study was approved by the NYU Grossman School of Medicine Institutional Review Board. A waiver of informed consent and a waiver of the Health Information Portability Privacy act were granted. The protocol was conducted in accordance to Declaration of Helsinki.

## RESULTS

Patients taking zinc sulfate in addition to hydroxychloroquine and azithromycin (n=411) and patients taking hydroxychloroquine and azithromycin alone (n=521) did not differ in age, race, sex, tobacco use or past medical history (Table 1). On hospital admission, vital signs differed by respiratory rate and baseline systolic blood pressure. The first laboratory measurements of inflammatory markers including white blood cell count, absolute neutrophil count, ferritin, D-dimer, creatine phosphokinase, creatinine, and C-reactive protein did not differ between groups. Patients treated with zinc sulfate had higher baseline absolute lymphocyte counts [median (IQR), zinc: 1 (0.7-1.3) vs. no zinc: 0.9 (0.6-1.3), p-value: 0.0180] while patients who did not receive zinc had higher baseline troponin [0.01 (0.01-0.02) vs. 0.015 (0.01-0.02), p-value: 0.0111] and procalcitonin [0.12 (0.05-0.25) vs 0.12 (0.06-0.43), p-value: 0.0493) (Table 1).

**Table 1:** Comparisons of baseline characteristics and hospital medications. Data are represented as median (IQR) or mean ± SD. Sample size is reported where it differed due to lab results not tested. P-values were calculated using 2-sided t-test for parametric variables and Mann Whitney U test for nonparametric continuous variables. Pearson χ^2^ test was used for categorical comparisons. *P* < .05 was deemed significant. Laboratory results represent the first measured value while hospitalized.

|  | **Zinc**<br>N=411 | **No Zinc**<br>N=521 | **P-value** |
| --- | --- | --- | --- |
| **Demographics** |  |  |  |
| Age | 63.19 ± 15.18 | 61.83 ± 15.97 | 0.0942 |
| Female Sex | 147 (35.7%) | 201 (38.6%) | 0.378 |
| Race |  |  | 0.428 |
| African American | 68 (16.5%) | 81 (15.5%) |  |
| White | 189 (46.0%) | 244 (46.8%) |  |
| Asian | 30 (7.3%) | 30 (5.8%) |  |
| Other | 97 (23.6%) | 142 (27.2%) |  |
| Multiracial/Unknown | 27 (6.6%) | 24 (4.6%) |  |
| **History** |  |  |  |
| Tobacco use |  |  | 0.142 |
| Never or Unknown | 306 (74.5%) | 382 (73.3%) |  |
| Former | 76 (18.5%) | 115 (22.1%) |  |
| Current | 29 (7.1%) | 24 (4.6%) |  |
| Any cardiovascular condition | 182 (44.3%) | 248 (47.6%) | 0.313 |
| Hypertension | 154 (37.5%) | 208 (39.9%) | 0.445 |
| Hyperlipidemia | 99 (24.1%) | 148 (28.4%) | 0.138 |
| Coronary Artery Disease | 36 (8.8%) | 41 (7.9%) | 0.624 |
| Heart Failure | 26 (6.3%) | 22 (4.2%) | 0.149 |
| Asthma or COPD | 50 (12.2%) | 56 (10.7%) | 0.499 |
| Diabetes | 105 (25.5%) | 130 (25.0%) | 0.835 |
| Malignancy | 23 (5.6%) | 33 (6.3%) | 0.638 |
| Transplant | 3 (0.7%) | 2 (0.4%) | 0.473 |
| Chronic Kidney Disease | 47 (11.4%) | 44 (8.4%) | 0.127 |
| BMI kg/m<sup>2</sup> | 29.17 (25.8-33.42) | 29.29 (25.77-33.2) | 0.8611 |
| **Admission Characteristics** |  |  |  |
| Oxygen saturation at presentation | 94 (91-96)* | 94 (91-96)** | 0.1729 |
| Respiratory Rate, respirations per minute | 20 (19-24) | 20 (18-24) | **0.0460** |
| Pulse, beats per minute | 97.66 ± 18.61 | 99.40 ± 19.82 | 0.0858 |
| Baseline Systolic BP, mmHg | 134.83 ± 20.84 | 132.41 ± 21.87 | **0.0435** |
| Baseline Diastolic BP, mmHg | 76.66 ± 12.62 | 76.59 ± 14.22 | 0.4670 |
| Temperature, degrees Celsius | 37.65 ± 0.82 | 37.72 ± 0.94 | 0.1354 |
| White blood cell count 10<sup>3</sup>/ul | 6.9 (5.1-9.0)<br>N=400 | 6.9 (5.1-9.3)<br>N=500 | 0.5994 |
| Absolute neutrophil count, 10<sup>3</sup>/ul | 5.15 (3.6-7.05)<br>N=388 | 5.4 (3.8-7.5)<br>N=488 | 0.0838 |
| Absolute lymphocyte count, 10<sup>3</sup>/ul | 1 (0.7-1.3)<br>N=388 | 0.9 (0.6-1.3)<br>N=482 | **0.0180** |
| Ferritin, ng/mL | 739 (379-1528)<br>N=397 | 658 (336.2-1279)<br>N=473 | 0.1304 |
| D-Dimer, ng/mL | 341 (214-565)<br>N=384 | 334 (215-587)<br>N=435 | 0.7531 |
| Troponin, ng/mL | 0.01 (0.01-0.02)<br>N=389 | 0.015 (0.01-0.02)<br>N=467 | **0.0111** |
| Creatine Phosphokinase, U/L | 140 (68-330)<br>N=343 | 151.5 (69.5-398.5)<br>N=344 | 0.4371 |
| Procalcitonin, ng/mL | 0.12 (0.05-0.25)<br>N=395 | 0.12 (0.06-0.43)<br>N=478 | **0.0493** |
| Creatinine, mg/dL | 0.97 (0.8-1.34)<br>N=400 | 0.99 (0.8-1.27)<br>N=499 | 0.4140 |
| C-Reactive Protein, mg/L | 104.95 (51.1-158.69)<br>N=398 | 108.13 (53-157.11)<br>N=480 | 0.9586 |
| **Medications recorded during hospitalization** |  |  |  |
| NSAID | 53 (12.9%) | 74 (14.2%) | 0.563 |
| Anticoagulant | 402 (97.8%) | 511 (98.1%) | 0.772 |
| ACE inhibitor or ARB | 138 (33.6%) | 175 (33.7%) | 0.997 |
| Beta Blocker | 91 (22.1%) | 132 (25.3%) | 0.256 |
| Calcium Channel Blocker | 89 (21.7%) | 104 (20.0%) | 0.527 |
| Corticosteroid | 40 (9.7%) | 47 (9.0%) | 0.711 |

\*measured on supplemental oxygen for 86.4%
\*\*measured on supplemental oxygen for 83.1%

In univariate analysis, the addition of zinc sulfate to hydroxychloroquine and azithromycin was not associated with a decrease in length of hospital stay, duration of mechanical ventilation, maximum oxygen flow rate, average oxygen flow rate, average fraction of inspired oxygen, or maximum fraction of inspired oxygen during hospitalization (Table 2). In bivariate logistic regression analysis, the addition of zinc sulfate was associated with decreased mortality or transition to hospice (OR 0.511, 95% CI 0.359-0.726), need for ICU (OR 0.545, 95% CI 0.362-0.821) and need for invasive ventilation (OR 0.562, 95% CI 0.354-0.891) (Table 3). However, after excluding all non-critically ill patients admitted to the intensive care unit, zinc sulfate no longer was found to be associated with a decrease in mortality (Table 3). Thus, this association was driven by patients who did not receive ICU care (OR 0.492, 95% CI 0.303-0.799). We also found that the addition of zinc sulfate was associated with likelihood of discharge to home in univariate analysis (OR 1.56, 95% CI 1.16-2.10) (Table 3). We performed a logistic regression model to account for the time-period when the addition of zinc sulfate to hydroxychloroquine plus azithromycin became utilized at NYULH. After adjusting for this date (March 25^th^), we still found an association for likelihood of discharge to home (OR 1.53, 95% CI 1.12-2.09) and decreased mortality or transition to hospice however the other associations were no longer significant (Table 4). The decrease in mortality or transition to hospice was most striking when considering only patients who were not admitted to the ICU (OR: 0.449, p-value: 0.002) (Table 4).

**Table 2:**
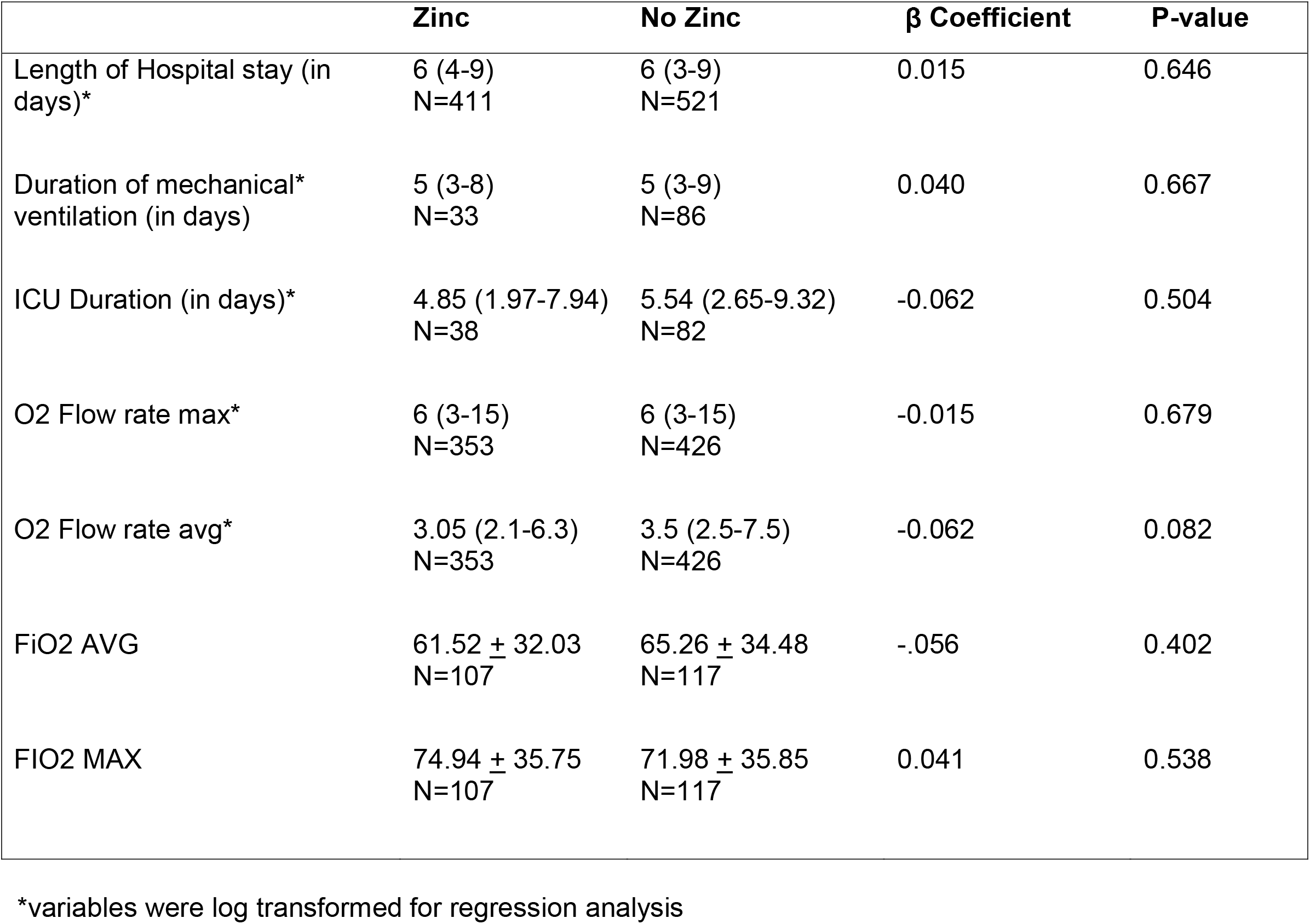
Comparisons of continuous hospital outcomes. Data are represented median (IQR) and as mean ± SD. Sample size is reported for each variable tested. β Coefficients and P-values were calculated using linear regression. N was specified for each comparison. *P* < .05 was deemed significant.

**Table 3:** Comparison of categorical hospital outcomes. Data are represented as N(%). P-values were calculated using logistic regression. *P* < .05 was deemed significant. N was specified for subgroup analyses.

|  | **Zinc** | **No Zinc** | **Odds Ratio** | **95% Confidence Interval** | **P-value** |
| --- | --- | --- | --- | --- | --- |
|  | N=411 | N=521 |  |  |  |
| Discharged home | 317 (77.1%) | 356 (68.3%) | 1.56 | 1.16-2.10 | **0.003** |
| Needed ICU | 38 (9.2%) | 82 (15.7%) | 0.545 | 0.362-0.821 | **0.004** |
| Needed Invasive Ventilation | 33 (8.0%) | 86 (16.5%) | 0.562 | 0.354-0.891 | **0.014** |
| Expired/Hospice | 54 (13.1%) | 119 (22.8%) | 0.511 | 0.359-0.726 | **<0.0001** |
| Expired/Hospice** | 28 (73.6%)<br>N=38 | 61 (74.4%)<br>N=82 | 0.964 | 0.401-2.31 | 0.934 |
| Expired/Hospice*** | 26 (6.9%)<br>N=373 | 58 (13.2%)<br>N=439 | 0.492 | 0.303-0.799 | **0.004** |

\*\*After excluding all non ICU patients
\*\*\*After excluding all ICU patients

**Table 4:** Adjusted comparison of categorical hospital outcomes. Data are represented as N(%). P-values were calculated using multivariate logistic regression adjusting for patient admission after March 25^th^ as a categorical variable. *P*□<□.05 was deemed significant. N was specified for subgroup analyses.

|  | **Zinc**<br>N=411 | **No Zinc**<br>N=521 | **Adjusted**<br>**Odds**<br>**Ratio** | **Adjusted**<br>**95%**<br>**Confidence**<br>**Interval** | **Adjusted**<br>**P-value** |
| --- | --- | --- | --- | --- | --- |
| Discharged home | 317 (77.1%) | 356 (68.3%) | 1.53 | 1.12-2.09 | **0.008** |
| Needed ICU | 38 (9.2%) | 82 (15.7%) | 0.733 | 0.471-1.14 | 0.168 |
| Needed Invasive Ventilation | 33 (8.0%) | 86 (16.5%) | 0.804 | 0.487-1.33 | 0.396 |
| Expired/Hospice | 54 (13.1%) | 119 (22.8%) | 0.559 | 0.385-0.811 | **0.002** |
| Expired/Hospice** | 28 (73.6%)<br>N=38 | 61 (74.4%)<br>N=82 | 1.03 | 0.404-2.64 | 0.947 |
| Expired/Hospice*** | 26 (6.9%)<br>N=373 | 58 (13.2%)<br>N=439 | 0.449 | 0.271-0.744 | **0.002** |

\*\*After excluding all non ICU patients
\*\*\*After excluding all ICU patients

## DISCUSSION

While practicing at the epicenter of the pandemic in the United States, we were faced with unprecedented challenges of adopting investigational therapies quickly into clinical practice. Initially, antiviral options at our institution consisted of clinician preference for either ritonavir/lopinavir or hydroxychloroquine plus azithromycin. After the findings of ritonavir/lopinavir in NEJM, we noticed an increase in the use of hydroxychloroquine plus azithromycin[16]. Our providers within the infectious diseases division, clinical pharmacy, and hospitalists discussed the use of zinc sulfate as an addition to hydroxychloroquine, based on the potential synergistic mechanism, and low risk of harm associated with this therapy.

To our knowledge, we provide the first *in vivo* evidence on the efficacy of zinc in COVID-19 patients. After adjusting for the timing of zinc sulfate treatment, the associations between zinc and the need for ICU and invasive ventilation were no longer significant but we did still observe a trend. This observation may be because patients with COVID-19 were initially sent to the ICU quicker, but as time went on and resources became more limited, clinicians began treating COVID-19 patients on general medicine floors for longer periods of time before escalating to the ICU. Future studies are needed to confirm or refute the hypothesis that the addition of zinc sulfate to a zinc ionophore such as hydroxychloroquine may reduce the need for ICU care in patients with COVID-19.

The main finding of this study is that after adjusting for the timing of zinc therapy, we found that the addition of zinc sulfate to hydroxychloroquine and azithromycin was found to associate with a decrease in mortality or transition to hospice among patients who did not require ICU level of care, but this association was not significant in patients who were treated in the ICU. This result may be reflective of the proposed mechanism of action of zinc sulfate in COVID-19. Zinc has been shown to reduce SARS-CoV RNA dependent RNA polymerase activity *in vitro* [13]. As such, zinc may have a role in preventing the virus from progressing to severe disease, but once the aberrant production of systemic immune mediators is initiated, known as the cytokine storm, the addition of zinc may no longer be effective [17]. Our findings suggest a potential therapeutic synergistic mechanism of zinc sulfate with hydroxychloroquine, if used early on in presentation with COVID-19. However, our findings do not suggest a prophylactic benefit of zinc sulfate in the absence of a zinc ionophore, despite interest in this therapy for prevention. A prophylactic strategy of zinc sulfate should be evaluated to help answer this question.

This study has several limitations. First, this was an observational retrospective analysis that could be impacted by confounding variables. This is well demonstrated by the analyses adjusting for the difference in timing between the patients who did not receive zinc and those who did. In addition, we only looked at patients taking hydroxychloroquine and azithromycin. We do not know whether the observed added benefit of zinc sulfate to hydroxychloroquine and azithromycin on mortality would have been seen in patients who took zinc sulfate alone or in combination with just one of those medications. We also do not have data on the time at which the patients included in the study initiated therapy with hydroxychloroquine, azithromycin, and zinc. Those drugs would have been started at the same time as a combination therapy, but the point in clinical disease at which patients received those medications could have differed between our two groups. Finally, the cohorts were identified based on medications ordered rather than confirmed administration, which may bias findings towards favoring equipoise between the two groups. In light of these limitations, this study should not be used to guide clinical practice. Rather, our observations support the initiation of future randomized clinical trials investigating zinc sulfate against COVID-19.

## Data Availability

Individual data not available for this study. For aggregate data, please contact corresponding author.

## Acknlowedgements

The authors thank Andrew Admon, Mary Grace Fitzmaurice, Brian Bosworth, Robert Cerfolio, Steven Chatfield, Thomas Doonan, Fritz Francois, Robert Grossman, Leora Horwitz, Juan Peralta, Katie Tobin, and Daniel Widawsky for their operational and technical support. We also thank the thousands of NYU Langone Health employees who have cared for these patients.

## References

1. Organization WH. Coronavirus Disease 2019 (COVID-19) Situation Report, 2020. Report No.: 46.

2. Huang C, Wang Y, Li X, et al. Clinical features of patients infected with 2019 novel coronavirus in Wuhan, China. The Lancet 2020; 395(10223): 497–506.

3. Walls AC, Park Y-J, Tortorici MA, Wall A, McGuire AT, Veesler D. Structure, Function, and Antigenicity of the SARS-CoV-2 Spike Glycoprotein. Cell 2020; 181(2): 281–92.e6.

4. Wu Z, McGoogan JM. Characteristics of and Important Lessons From the Coronavirus Disease 2019 (COVID-19) Outbreak in China: Summary of a Report of 72⍰314 Cases From the Chinese Center for Disease Control and Prevention. JAMA 2020; 323(13): 1239–42.

5. Zhou F, Yu T, Du R, et al. Clinical course and risk factors for mortality of adult inpatients with COVID-19 in Wuhan, China: a retrospective cohort study. The Lancet 2020; 395(10229): 1054–62.

6. Zhou P, Yang X-L, Wang X-G, et al. A pneumonia outbreak associated with a new coronavirus of probable bat origin. Nature 2020; 579(7798): 270–3.

7. Savarino A, Boelaert JR, Cassone A, Majori G, Cauda R. Effects of chloroquine on viral infections: an old drug against today’s diseases. The Lancet Infectious Diseases 2003; 3(11): 722–7.

8. Vincent MJ, Bergeron E, Benjannet S, et al. Chloroquine is a potent inhibitor of SARS coronavirus infection and spread. Virology Journal 2005; 2(1): 69.

9. Yao X, Ye F, Zhang M, et al. In Vitro Antiviral Activity and Projection of Optimized Dosing Design of Hydroxychloroquine for the Treatment of Severe Acute Respiratory Syndrome Coronavirus 2 (SARS-CoV-2). Clin Infect Dis 2020: ciaa237.

10. Gautret P, Lagier J-C, Parola P, et al. Hydroxychloroquine and azithromycin as a treatment of COVID-19: results of an open-label non-randomized clinical trial. International Journal of Antimicrobial Agents 2020: 105949.

11. Magagnoli J, Narendran S, Pereira F, et al. Outcomes of hydroxychloroquine usage in United States veterans hospitalized with Covid-19. medRxiv 2020: 2020.04.16.20065920.

12. Molina JM, Delaugerre C, Le Goff J, et al. No evidence of rapid antiviral clearance or clinical benefit with the combination of hydroxychloroquine and azithromycin in patients with severe COVID-19 infection. Médecine et Maladies Infectieuses 2020.

13. te Velthuis AJW, van den Worm SHE, Sims AC, Baric RS, Snijder EJ, van Hemert MJ. Zn2+ Inhibits Coronavirus and Arterivirus RNA Polymerase Activity In Vitro and Zinc Ionophores Block the Replication of These Viruses in Cell Culture. PLOS Pathogens 2010; 6(11): e1001176.

14. Maret W. Zinc in Cellular Regulation: The Nature and Significance of “Zinc Signals”. Int J Mol Sci 2017; 18(11): 2285.

15. Xue J, Moyer A, Peng B, Wu J, Hannafon BN, Ding W-Q. Chloroquine is a zinc ionophore. PloS one 2014; 9(10): e109180-e.

16. Cao B, Wang Y, Wen D, et al. A Trial of Lopinavir–Ritonavir in Adults Hospitalized with Severe Covid-19. New England Journal of Medicine 2020.

17. Li X, Geng M, Peng Y, Meng L, Lu S. Molecular immune pathogenesis and diagnosis of COVID-19. Journal of Pharmaceutical Analysis 2020.

